# COVID-19 Risk Stratification and Mortality Prediction in Hospitalized Indian Patients

**DOI:** 10.1101/2020.12.19.20248524

**Authors:** Shanmukh Alle, Samreen Siddiqui, Akshay Kanakan, Akshit Garg, Akshaya Karthikeyan, Neha Mishra, Swati Waghdhare, Akansha Tyagi, Bansidhar Tarai, Pranjal Pratim Hazarika, Poonam Das, Sandeep Budhiraja, Vivek Nangia, Arun Dewan, Ramanathan Sethuraman, C. Subramanian, Mashrin Srivastava, Avinash Chakravarthi, Johnny Jacob, Madhuri Namagiri, Varma Konala, Debasish Dash, Sujeet Jha, Rajesh Pandey, Anurag Agrawal, P. K. Vinod, U. Deva Priyakumar

## Abstract

The clinical course of coronavirus disease 2019 (COVID-19) infection is highly variable with the vast majority recovering uneventfully but a small fraction progressing to severe disease and death. Appropriate and timely supportive care can reduce mortality and it is critical to evolve better patient risk stratification based on simple clinical data, so as to perform effective triage during strains on the healthcare infrastructure. This study presents risk stratification and mortality prediction models based on usual clinical data from 544 COVID-19 patients from New Delhi, India using machine learning methods. An XGboost classifier yielded the best performance on risk stratification (F1 score of 0.81). A logistic regression model yielded the best performance on mortality prediction (F1 score of 0.71). Significant biomarkers for predicting risk and mortality were identified. Examination of the data in comparison to a similar dataset with a Wuhan cohort of 375 patients was undertaken to understand the much lower mortality rates in India and the possible reasons thereof. The comparison indicated higher survival rate in the Delhi cohort even when patients had similar parameters as the Wuhan patients who died. Steroid administration was very frequent in Delhi patients, especially in surviving patients whose biomarkers indicated severe disease. This study helps in identifying the high-risk patient population and suggests treatment protocols that may be useful in countries with high mortality rates.

## Introduction

The World Health Organization (WHO) declared the outbreak of coronavirus disease 2019 (COVID-19) as a global health emergency of international concern. Originating in Wuhan, China, the disease has spread to the rest of the world. As of December 17, 2020, WHO states that about 10 million confirmed cases of COVID-19 have been detected in India alone, making the number of cases in the country 15% of the total cases worldwide and the second largest affected nation after the United States. Due to the sudden spike in the number of cases, healthcare systems across the world including India’s are under tremendous pressure for making tough decisions in resource allocation among affected patients. Early risk stratification through identification of key biomarkers is essential because it helps in the understanding of the relative severity among infected patients and hence guides decisions in the scare medical resource setting.

COVID-19 is a highly contagious respiratory infection with symptoms that include fever, dry cough, nasal congestion and breathing difficulties [1, 2]. In more severe cases, it can cause pneumonia, severe acute respiratory syndrome, cardiac arrest, sepsis, kidney failure and death [3, 4]. WHO classifies the risk into the following categories: critical, severe, and moderate/mild. By definition, critical patients require ventilation, severe patients require supplemental oxygen, moderate patients have pneumonia but do not require oxygen, and mild patients only have upper respiratory tract disease. The cause of death is generally respiratory failure, but few deaths have been caused by multiple organ failure (MOF) or chronic comorbidities [2, 5]. Those at a higher risk are the elderly and people with comorbidities, such as cardiovascular diseases and diabetes [6, 7]. However, symptoms at onset are relatively mild and a significant proportion of patients do not show apparent symptoms prior to the development of respiratory failure [2, 5]. Clinically, this makes it difficult to predict the progression of severity in patients until respiratory failure develops. Early risk prediction and effective treatment can reduce mortality and morbidity as well as relieve resource shortages [8]. Artificial intelligence based solutions may help in clinical decision-making by providing predictions that are accurate, fast, and interpretable. Recent studies have used various machine learning algorithms for analysing COVID-19 patients’ clinical data and providing disease prognosis [9, 10]. Hao et al. [11] examined COVID-19 patients admitted in Massachusetts to predict level-of-care requirements based on clinical and laboratory data. They compared machine learning algorithms (such as XGBoost, Random Forests, SVM, and Logistic Regression) and predicted the need for hospitalization, ICU care, and mechanical ventilation. The most effective features for hospitalization were vital signs, age, BMI, dyspnea, and comorbidities. Opacities on chest imaging, age, admission vital signs and symptoms, male gender, admission laboratory results, and diabetes were the most effective risk factors for ICU admission and mechanical ventilation.

Xie et al. [12] used multivariable logistic regression for the classification task through identifying SpO2, Lymphocyte Count, Age and Lactate dehydrogenase (LDH) as the set of important features. A nomogram was created based on these features to deliver the probability of mortality. Ji et al. [13] built a scoring model, named as CALL, for prediction of progression risk in COVID-19 patients from Chinese hospitals. They used Multivariate Cox regression to identify risk factors associated with progression, which were then incorporated into a nomogram for establishing a prediction scoring model. Comorbidity, older age, lower lymphocyte count, and higher lactate dehydrogenase were found to be independent high-risk factors for COVID-19 progression. Yan et al. proposed an interpretable mortality prediction model for COVID-19 patients [14]. They analysed blood samples of 485 patients from Wuhan, China, and created a clinically operable single tree through XGBoost. The model used three crucial features Lactate Dehydrogenase (LDH), lymphocyte (%) and C-Reactive Protein (CRP). The decision rules with the three features and their thresholds were devised recursively. This provided an interpretable machine learning solution with at least 90% accuracy. Karthikeyan et al. [15] analysed the same dataset through comparing various machine learning algorithms. XGBoost feature selection and neural network classification yielded the best performance with the important biomarkers selected as neutrophil (%), lymphocyte (%), LDH, CRP and age. However, no detailed studies on risk stratification have not been done on Indian cohorts.

Most machine learning based risk stratification and mortality prediction algorithms analysed patients from China or the United States of America. Studies have suggested that the virus has different strains around the globe due to mutations [16–20]. Moreover, the physiologic response to the virus and the eventual course of disease depends on regional factors such as population characteristics and hospital practices. Hence, the studies are not universally applicable and it is critical to examine cohorts from India to aid the Indian healthcare systems. In this study, patients with confirmed COVID-19 infection from MAX group of hospitals in New Delhi, India were examined to identify the key features affecting severity and mortality. The machine learning models built using these key features aid in risk stratification and mortality prediction. The mortality rate in the Indian population is low compared to China and other countries. Hence a comprehensive comparison between the cohorts from New Delhi and Wuhan has been done, and analysis with respect to treatment protocols were explored to identify possible factors for such a difference [14].

## Methodology

### Data acquisition and Participants

The data in this study was collected from patients with confirmed diagnosis of COVID-19 at MAX group of Hospitals in New Delhi, India between June 3rd and October 23rd, 2020. The patient records were collected and anonymized at the data warehouse of CSIR-IGIB. A total of 544 patients with a clear final outcome were considered in our study. Among these, diagnostic lab reports were available as a time series of test results. The data collected contains 357 distinct parameters (or biomarkers) that include vitals, symptoms, co morbid conditions and lab reports from 161 different tests along with the medicines administered for treatment. Multiple tests were recorded for each patient during their stay at the hospital, varying from 1 to 134 records per patient.

### Risk Stratification and Statistical Analysis

Patients were categorized into risk levels-based on the severity of their condition during their stay at the hospital. Although a description of the severity of each patient is not available, since COVID-19 is a respiratory disorder that effects the lungs, the amount of care provided to a patient with respect to respiratory support was considered as an indicator of severity. Considering the size of the dataset and the levels of respiratory support provided all the patient were categorized into two levels mild and severe where all patients who died or who were under some form of respiratory support or whose condition was specifically mentioned to be severe were categorized into severe/high risk group and all the remaining patients were put under mild/low risk group. The resulting dataset follows the data distribution as shown in Table 1.

**Table 1.**
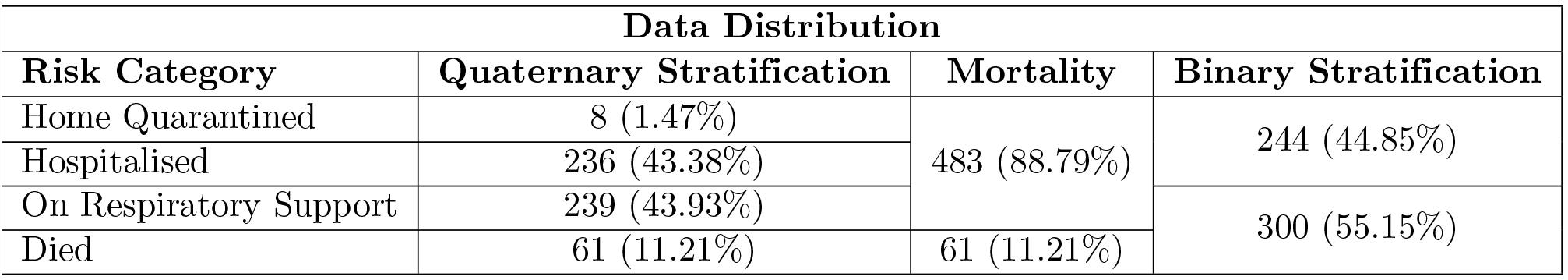
Distribution of the number of patients across various classes.

The 15 most frequent tests corresponding to 38 biomarkers were selected for analysis based on the availability of data. Five biomarkers WBC count, neutrophil lymphocyte ratio (NLR), lymphocyte monocyte ration (LMR), neutrophil monocyte ratio (NMR), platelet to lymphocyte ratio (PLR) were manually calculated from various blood cell counts available owing their reported importance in predicting mortality due to COVID [21, 22]. 209 unique co-morbid conditions were observed in patients in our study. To analyse them without exploding the number of features and to avoid an increase in chances of over fitting due to increase in dimensionality, we grouped all the encountered co-morbid conditions into 7 groups based on the area or organ that the condition effects as shown in Table S1. Four more groups: diabetes, hypertension, hyperlipidaemia, and cancer were considered due to their reported importance in mortality risk due to COVID-19 [7]. This information was encoded into 11 binary features, each representing one group where a sample assumes a value one if the patient has one or more co-morbid conditions that fall into that group. To incorporate and analyse the effects of medical prescriptions the information regarding prescription of steroids and antiviral drugs was encoded into two binary features.

This leads to 70 unique parameters measured which include 11 grouped co-morbid conditions, 14 clinical parameters, 2 RT-PCR genomic parameters and 43 lab test results. An exhaustive list of categorical parameters can be found in Table S1 and continuous parameters can be found in Table S2. To evaluate the significance of each parameter considered for risk stratification and mortality prediction, we calculated the p value using the Chi-Squared test [23] for the categorical features and using the ANOVA f-value test for the continuous features.

### Comparison with Wuhan cohort

In our study, we evaluated how machine learning models trained on non-Indian cohorts perform in predicting mortality on the Indian cohort. We used the best performing model reported by Karthikeyan et.al [15] for predicting mortality using data from Wuhan, China [14] to examine its applicability on the Indian cohort. The Wuhan cohort comprises of data collected from 375 patients who were admitted to Tongji Hospital, Wuhan. The model evaluated is a neural network trained to predict mortality from CRP, LDH, neutrophil (%) lymphocyte(%) and age. For predicting mortality in Indian Cohort using the same model, we selected 3092 datapoints where at least 3 of the required 5 features were present. KNN imputation was done to take care of the missing features.

We also explored the differences between Wuhan and New Delhi cohorts in key biomarkers across survivors and the dead [14, 15, 24]. We choose mortality as the indicator for comparison as it does not depend on subjective labelling. The feature density histograms were analyzed to examine the variations in biological parameters across survivors and the dead between cohorts of Wuhan and Delhi. The Kolmogorov–Smirnov test (K-S test) [25] was used to analyze variations in the density distributions of the important biomarkers between both classes across cohorts. The K-S test is a non-parametric test that quantifies the distance between the empirical distributions of samples sampled from two distributions.

### Machine Learning Pipeline

Figure 1 depicts the overall pipeline used in this study for performing the risk stratification and mortality prediction tasks. We compared several machine learning algorithms namely XGBoost, random forests, Support Vector Machine (SVM) and logistic regression for evaluating their predictive performance. A detailed account of the step-by-step procedure is presented in the following sections.

**Figure 1.**
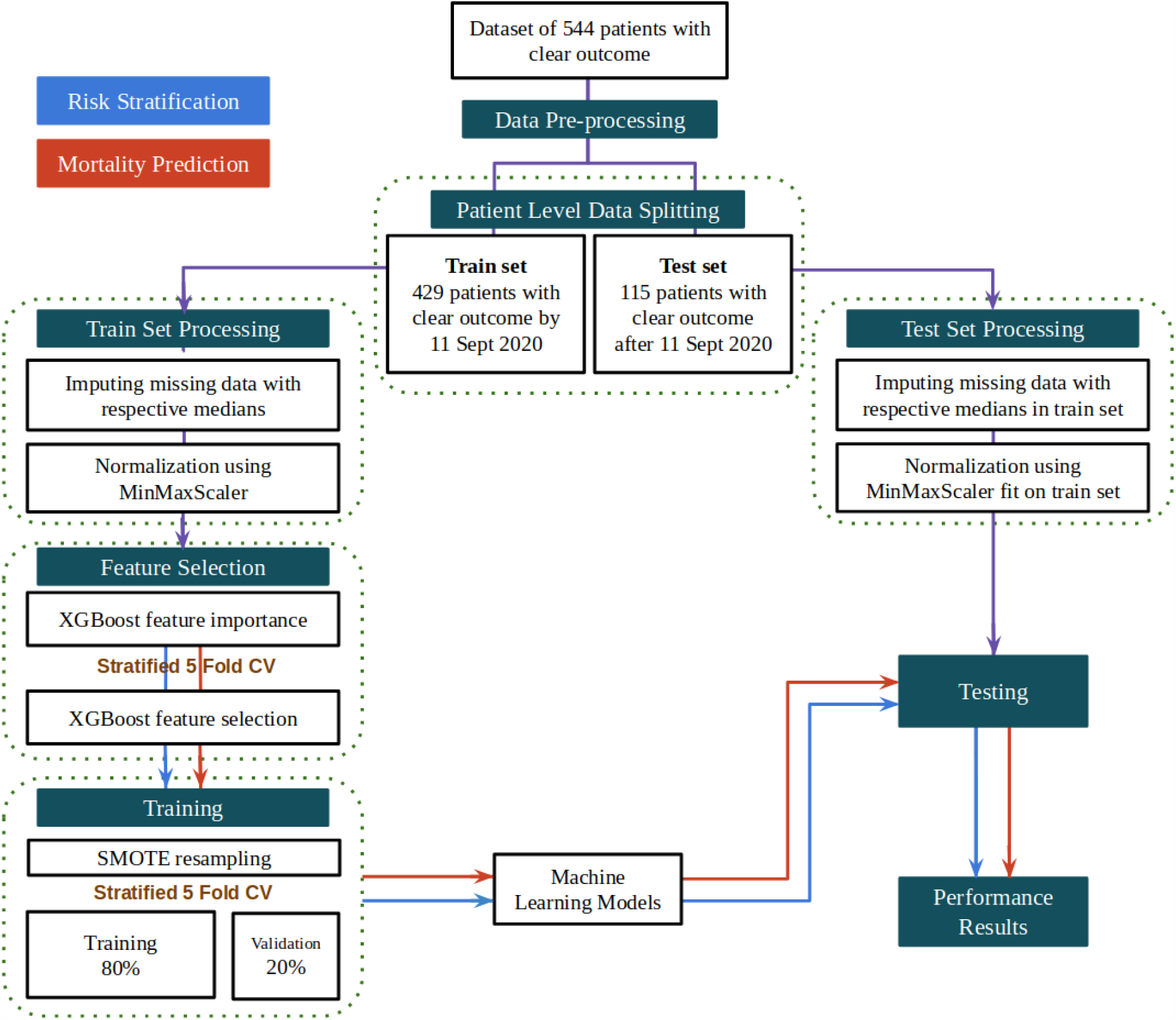
Machine learning pipeline for the development of the risk stratification and mortality prediction tasks.

### Data Pre-processing

For each patient in the dataset, there were multiple lab test results recorded on different days before the outcome. We have considered each individual recorded test result as a unique data point for training and testing as has been done before [14, 15]. Each sample has a dimensionality equal to the number of unique parameters measured across all lab tests considered for the analysis. The values in a sample are filled in with the test results that a particular sample represents and the rest of the values are left empty. These parameter values that are left empty are imputed with the nearest value of the parameter from the patient’s past test results. Some samples may still have missing parameters if a patient does not undergo a particular test. Such missing values are imputed with the median of the respective parameter across the train set. Patient demographics and vitals data were recorded once per patient and were added to each sample where they are kept the same for all the samples of a particular patient. This leads to 15648 samples from 544 patients where each sample contains 70 unique parameters. The resulting dataset follows the data distribution as shown in Figure 2.

**Figure 2.**
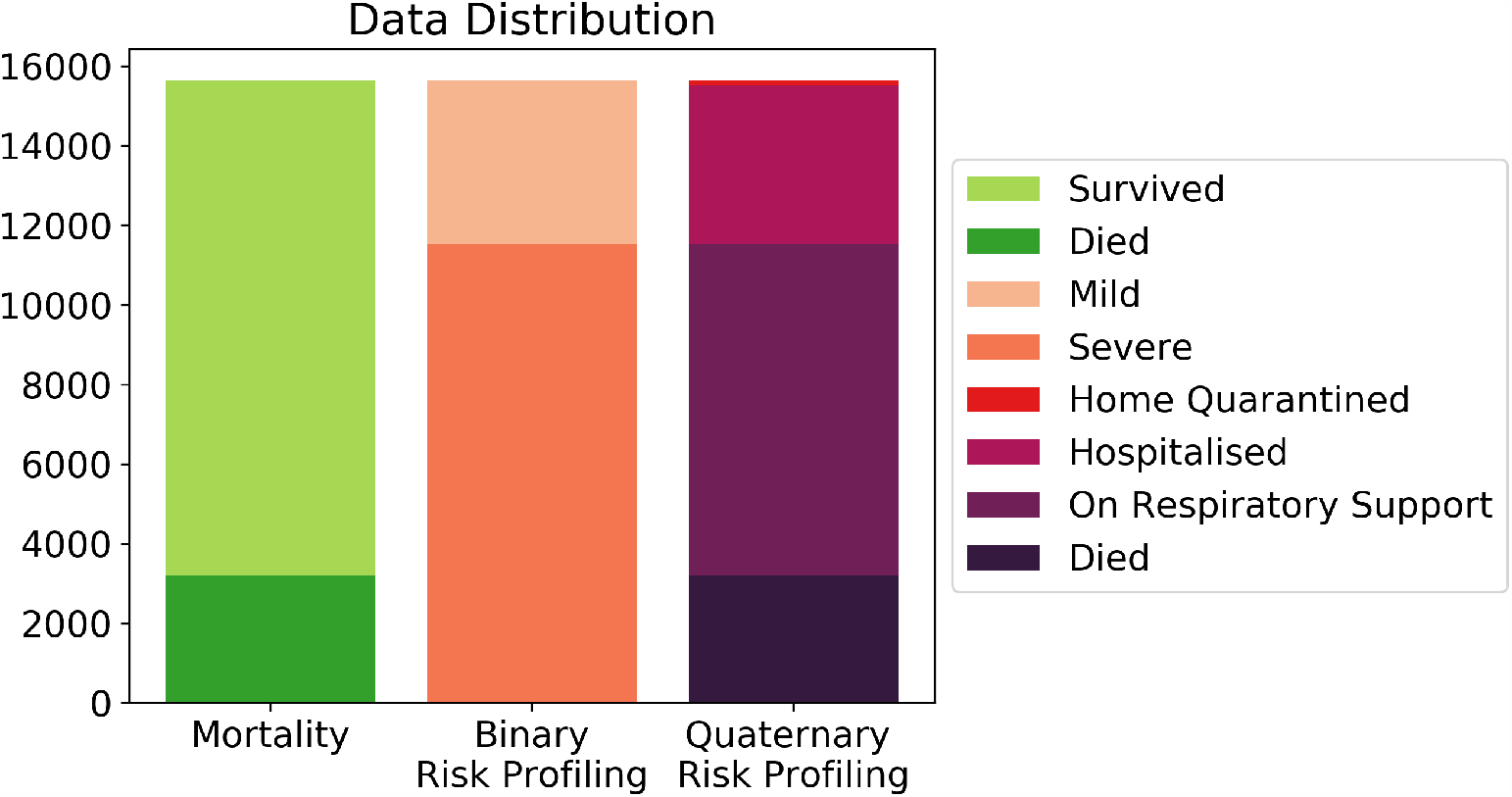
Distribution of the data with respect to mortality, binary risk profiles and quarternary risk profiles.

To build and validate machine learning models we split patients with respect to the day of outcome. 429 patients with clear outcome by 11 September 2020 were considered for model development, and the remaining 115 patients were considered as a part of a holdout test set. This method of splitting is adopted as models developed will be used to aid future patients where it is known that the COVID-19 and responses of its infected patients may change with time [16–19]. The day wise distribution of samples in both the train and test sets for risk stratification and mortality prediction is shown in Figures S1 and S2, respectively.

### Feature Selection

Among the 70 features chosen for analysis, selecting the most influential biomarkers for risk stratification and mortality prediction by eliminating redundant or unimportant parameters is crucial to avoid over-fitting when the size of the dataset is small. Moreover, a lower number of features would mean cheaper and faster tests for efficient risk profiling given the high influx of patients on a daily basis and subsequently increased efficiency of the decision-making process of the healthcare systems.

The relative importances provided by an XGBoost classifier fit on the training data for a particular task is used as the measure of importance for selecting features. XGBoost is a powerful decision-tree-based ensemble algorithm that uses a gradient boosting framework and estimates features that are the most discriminative of model outcomes [26]. The relative importance of each feature is determined by its accumulated use in each decision step in each tree of the ensemble.

The number of features to utilize for model training was obtained by iteratively training an XGboost model on a collection of the top K most important features while increasing K by 1 in each iteration. The collection of features that achieved the best performance for 5-fold cross validation on the training set was considered as the set of key features to train the final models. The feature importances were obtained separately for the binary risk stratification and mortality prediction models. The classification performance for selecting the optimal set of features is evaluated using AUC score for risk stratification and average precision score for mortality prediction. Average precision score is used for mortality prediction due to the heavy imbalance of samples representing fatal cases in mortality prediction.

### Training

After obtaining the collection of important features, duplicates that arose due to the elimination of less important features were removed from the train set. The set was then normalized to a range of 0-1 using min-max scaler to avoid any biases due to differences in scales across parameters. The train set was then resampled using the SMOTE algorithm to reduce bias that may arise due to the class imbalance observed. The SMOTE algorithm was chosen to generate synthetic samples of the minority class due to its good performance. Various algorithms were trained and compared on the resampled dataset to classify the samples depending on the task, either risk stratification or mortality prediction, with their respective feature set. We also built another set of models trained on only patient vitals to gauge the prediction performance that can be achieved with data acquired before blood test results.

### Testing

The hold out test data of 115 patients was normalized with min-max scaler to a range of 0-1 using the min-max statistics obtained from the training set. Then the models built were evaluated on the test set. We report the F1-scores of the algorithms as the mean and standard deviation of performance of trained models from 5-fold cross validation on the test set. F1-score is preferred over AUC and accuracy as it is better in measuring performance when data imbalance exists. The model achieving the best performance was then tested and analysed on the set of samples corresponding to each individual day for a period of 14 days before the final outcome to observe relevant trends.

### Evaluation Metrics

The following metrics were recorded to assess the predictive performance of the supervised models. Formulae for the calculation of all metrics are given below. Here, TP, TN, FP, and FN stand for true positive, true negative, false positive and false negative rates, respectively.

#### AUC (Area under ROC curve)

AUC measures the area under the receiving operator characteristic (ROC) curve, which plots true positive rate against false positive rate. AUC is also commonly used in situations where the data has imbalanced classes, as the ROC measures performance over many different thresholds.

- True Positive Rate (TPR):This measures how often the model predicts that a patient will survive when the person survives.

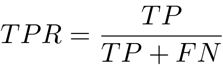
- False Positive Rate (FPR): This measures how often the model predicts that a patient survives when the person actually does not survive:

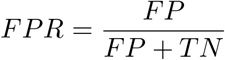

#### F1 Score

The F1 score measures the harmonic mean of precision of recall and is often preferred to accuracy when the data has imbalanced classes:

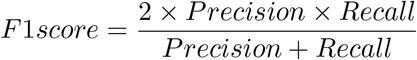

where,

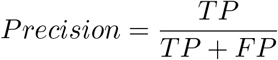

and,

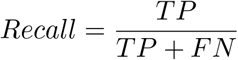

## Results

### Patient Characteristics

A comparison of different clinical features between low and high-risk patients was carried out. Tables S1-S4 show the differences in categorical and continuous features between high and low risk groups, and between survivors and the dead. The KS test showed that none of the continuous features followed a normal distribution and hence the medians and interquartile ranges are reported. The patients’ age ranged between the age of 9 and 98 years with the median age 58 (48-66) years. The median age for the high-risk patients was 61 (53-68) years while for the low-risk patients it was 53 (41-64) years. Out of the 544 patients, 164 (30.15%) were females while 380 (69.85%) were males. The blood clotting (D-Dimer, Ferritin), inflammation (CRP, LDH) and immune features (NLR, LMR, NMR, PLR and IL6) were significantly different for the low and high-risk groups. However, a significant overlap was observed in most of the parameters both when comparing the high-risk vs. low-risk and survived vs. dead categories precluding the possibility of the development of simple classification models.

### Indian vs. Wuhan cohort

Machine learning models for predicting mortality based on patients’ blood parameters have been reported, however, it is necessary to ascertain whether these models are generalizable. Karthikeyan et.al. [15] built a neural network that predicted mortality in Wuhan cohort with an accuracy of 96.5%*±*0.6% using only five parameters, age, lymphocyte (%), neutrophil (%), LDH and CRP. The same model when tested on the Indian cohort (current dataset) predicted mortality with an accuracy of only 58%. The drop in performance of the model when tested on the Indian group shows that there is a significant difference between the two cohorts. Figure 3 demonstrates that the Neural Net was performing much better in identifying the patients who died (precision 84.85%) over those who survived (precision 49.54%). This suggests that the patients who were expected to die based on the findings from Wuhan data were actually surviving in the Indian Cohort.

**Figure 3.**
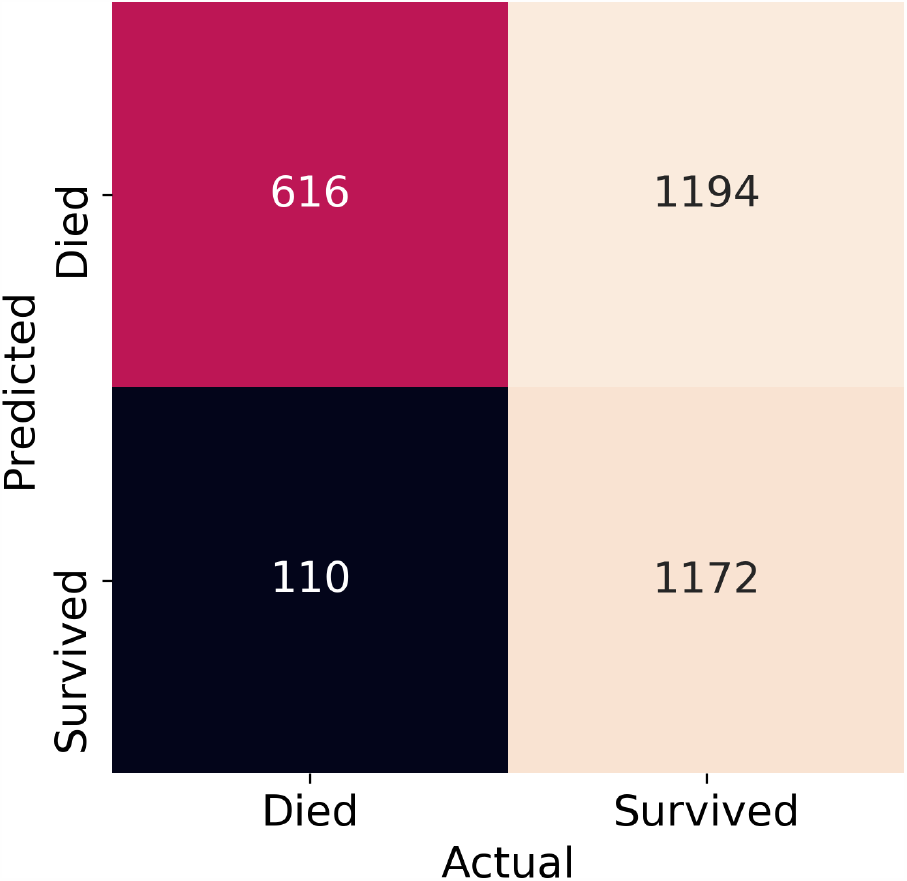
Confusion matrix of neural net net trained on Wuhan data and tested on Indian data

To understand the difference between cohorts, we compared the feature density histograms of Indian and Wuhan cohorts (Figure 4). It was observed that survival of patients with LDH in the range 500-1000 is much higher in Delhi compared to Wuhan. It can also be observed that there are almost no survivors with an LDH value greater than 800 in the Wuhan cohort while patients with LDH values of even about 1000 have survived in Delhi Cohort. The survivability of patients with CRP greater than 50 is higher in the Indian cohort compared to Wuhan. Similar conclusions can be drawn with Indian patients having relatively lower lymphocyte (%) and higher neutrophil (%). This is interesting as the likelihood of survival with higher neutrophil (%) or lower lymphocyte (%) is much lower [27].

**Figure 4.**
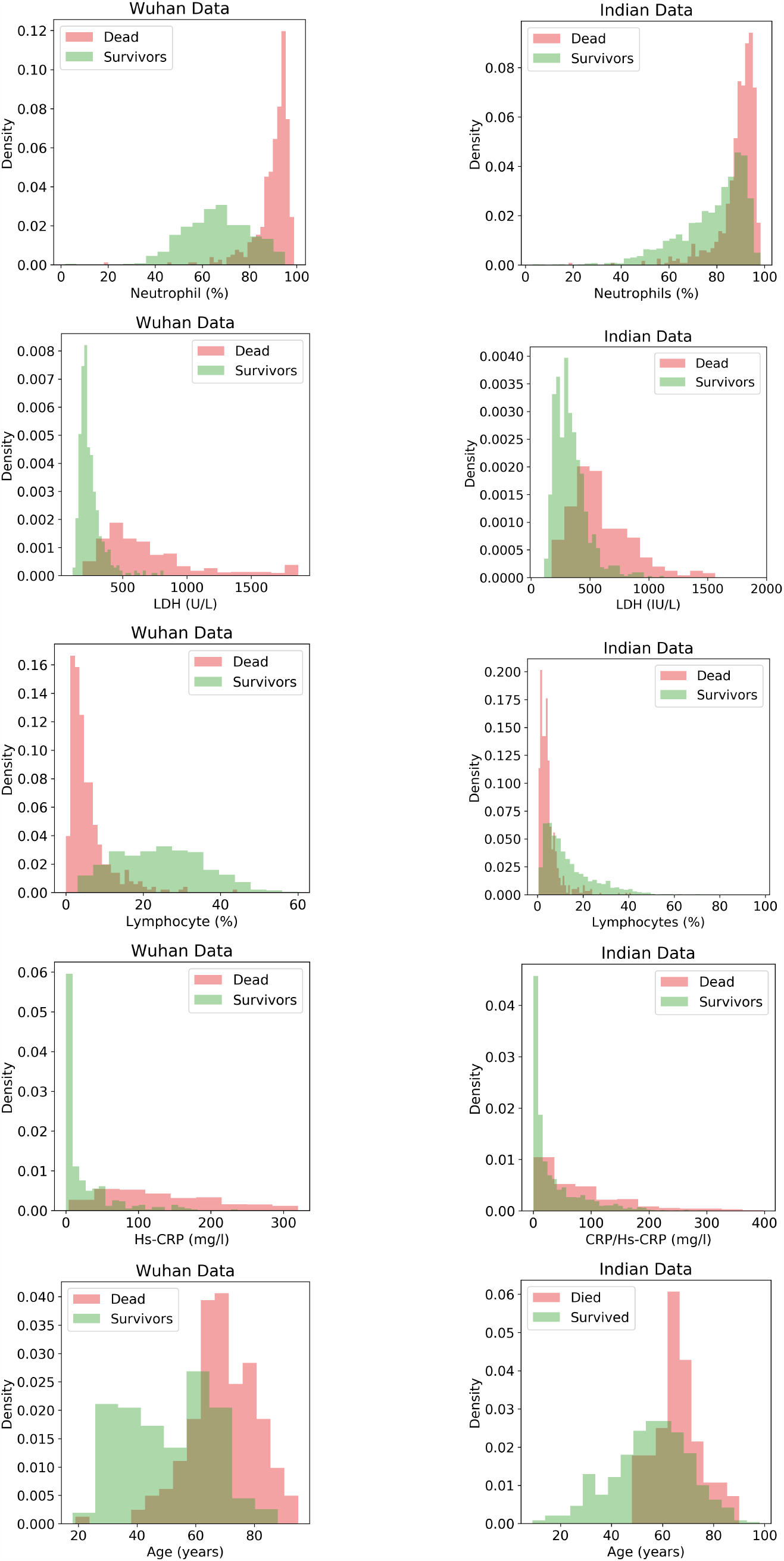
Comparison of the the normalized histogram plots of important features useful for predicting mortality from Wuhan and Indian Cohorts.

Figure 5 shows various matrices with two sample K-S statistics that measure pairwise distances between distributions of important biomarkers of survivors and the dead across Indian and Wuhan cohorts. It is observed that the distance between distributions of the Indian Recovered (IR) and Indian Dead (ID) is significantly lower compared to the distance between the distributions of the Wuhan Recovered (WR) and Wuhan Dead (WD) for all the five biomarkers. This is mainly due to the differences between distributions of recovered across Delhi and Wuhan as the distance between the cohorts of the dead is low and the distance between cohorts of the recovered is high. From this, it is evident that many Indian patients who were at high risk of death according to the insights from other cohorts have survived.

**Figure 5.**
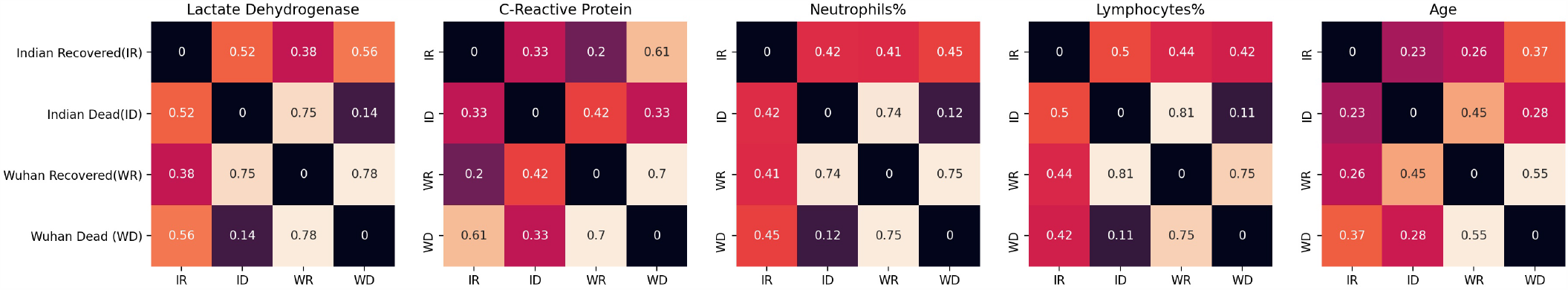
Pairwise distances between distributions of important features across the Indian vs. Wuhan survived and dead classes. Distance values were calculated through Kolmogorov–Smirnov test.

It is observed that the distance between WD and ID distributions is low, especially in neutrophils (%), LDH and lymphocytes (%). The high survivability of patients with extreme neutrophil and lymphocyte percentages is consistent with the lower mortality rates observed in the Indian population compared to several other countries. Identification of possible reasons for such a phenomenon would enable minimization of mortality in other countries as well.

### Risk Stratification

XGboost was used to rank features based on the contribution of each features to the performance in risk stratification. Figure S3 shows the list of the top 25 features sorted in descending order with respect to their relative importance in risk stratification. The 11 features that were selected to train the models in the order of their importance are absolute neutrophil count, LDH, lymphocyte (%), neutrophil(%), record of diabetes comorbidity, ferritin, INR, interleukin-6(IL-6), oxygen saturation level, absolute eosinophil count and packed cell volume. Figure S4 shows the density distributions for the top 4 features identified.

Comparison of the performance of various algorithms showed XGboost algorithm to perform the best with an F1-score of 0.810*±*0.01 as seen in Figure 6. The model also yielded better AUC (0.833*±*0.01) and average precision (0.891*±*0.01) (Table S5). The confusion matrix of predictions from an XGboost model trained on the entire train set is shown in Figure S5. We also evaluated how the performance of model changes with days to outcome, where the day of outcome is either the day of discharge from the hospital or the day of death. Figure 7 shows that the performance of the risk stratification model decreases as the samples approach the day of outcome. This suggests that the feature difference between low risk and some high-risk patients who are recovering is decreasing towards the day of outcome. However the performance of the mortality prediction model increases towards the day of outcome. Hence, selective use of these two models depending on the number of days from infection may be effective.

**Figure 6.**
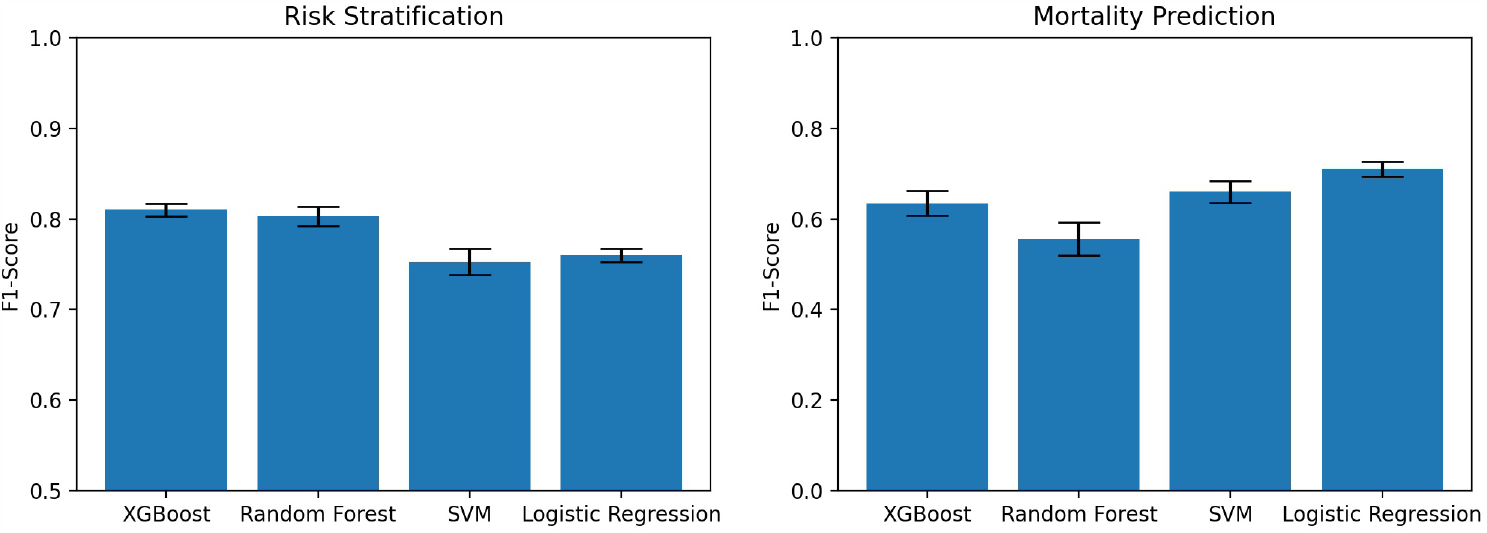
Comparison of F1 scores for various machine learning models that use patient vitals and lab test results.

**Figure 7.**
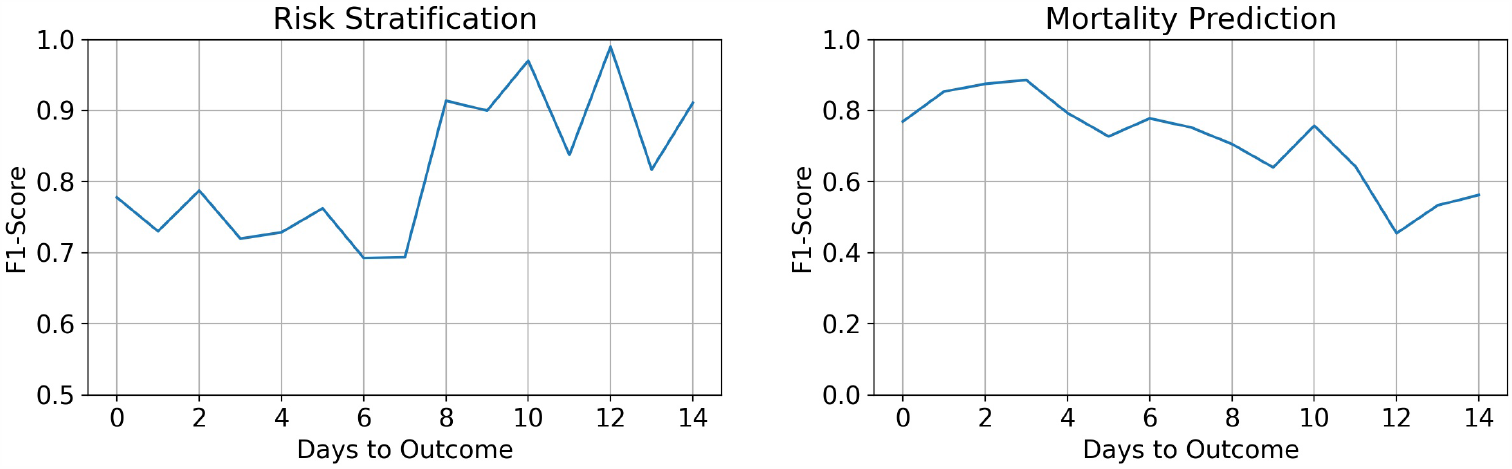
Performance of the ML models with respect to number of days to outcome.

Further, we trained and evaluated models with only patient vitals, comorbidities, and medication information to evaluate the predictive performance that can be achieved without lab test results. Figure S6 shows the F1 scores of various models that were built to use only these patient information. The random forests algorithm performed the best with an F1 score of 0.76±0.02. The important features selected were administration of steroids, oxygen saturation levels, record of diabetes, thyroid problems, presence of any other comorbidities, weight, temperature, respiration rate, hypertension, and BMI.

### Mortality Prediction

Figure S7 shows the top 25 features sorted in descending order with respect to their relative importance in mortality prediction. The 9 features that were selected to obtain the results in the order of their importance are D-Dimer, Ferritin, Lymphocyte (%), Neutrophil to Lymphocyte ratio (NLR), WBC, Trop I, INR, IL-6 and LDH. Figure S8 shows the density distributions for the top 4 features identified.

Logistic regression model performed the best with an F1-score of 0.710*±*0.02 as seen in Figure 6. The model also yielded better AUC (0.927*±*0.01) and average precision (0.801*±*0.02) (Table S6). The performance of the model increases as the samples approach the day of outcome as seen in Figure 7. We trained and evaluated models with only patient vitals, comorbidities, and medication information to evaluate the predictive performance that can be achieved with data excluding lab test results. Figure S6 shows the F1-scores of various models that were built using the selected patient information. SVM performed the best with an F1 score of only 0.34*±*0.03. The important features selected were hypertension, record of any comorbidities related to liver, record of cancer, oxygen saturation, administration of antivirals and respiration rate.

### Role of Steroids

As a part of the study, we also compared the differences in neutrophil and lymphocyte percentages across patients who were administered steroids and patients who were not to understand if the treatment protocols followed in India medical systems has an effect on the lower mortality rates. Of the 544 patients involved in the study 338 (62.13%) patients were administered steroids. It was observed that Methylprednisolone was the most widely administered steroid that was given to 262 different patients, followed by Dexamethasone given to 89 patients. Prednisolone was administered to 11 patients while Hydrocortisone and Triamcinolone were given to only one patient. It is to be noted that there were instances where a single patient was administered with more than one of these. Figure 8 shows the density histograms of neutrophil and lymphocyte percentages for survivors and mild patients. It is observed that a higher proportion of the survivors and mild patients who were administered steroids had extreme neutrophil and lymphocyte percentages indicating that administration of steroids may have had an impact in patient outcome.

**Figure 8.**
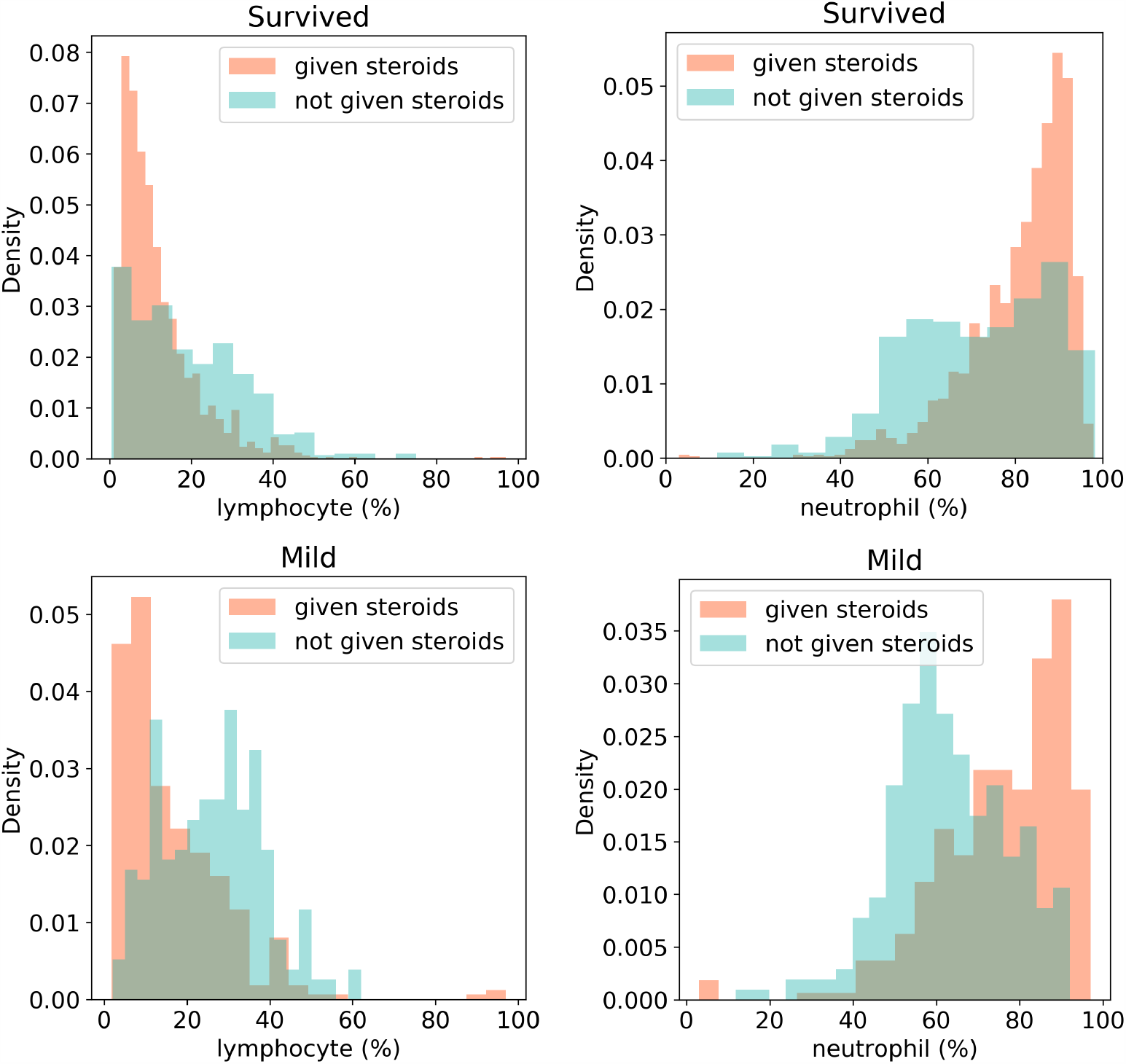
Distribution plots for lymphocytes (%) and neutrophil (%) for the patients who survived and were administered steroids vs those who survived and were not administered steroids and patients who had mild severity and were administered steroids vs patients who had mild severity and were not administered steroids.

## Discussion

COVID-19 has spread around the globe and the need for fast and effective resource allocation is urgent, but very few studies have examined Indian cohorts. Studies show the effect of regional factors such as patient characteristics and method of treatment on their response to the infection. Hence, it is critical to analyse Indian patients to aid the healthcare systems of the second highest COVID-19 hit nation in the world. In this study, we analysed 15648 samples of 544 patients, with confirmed diagnosis of COVID-19, at MAX group of hospitals in New Delhi, India. Each sample contains 70 unique parameters including the grouped comorbid conditions, patient vitals, patient demographic information, and lab test results. We found that existing mortality prediction models trained on Wuhan cohort cannot be directly used for mortality prediction on the Indian cohort due to cohort specific differences in response to COVID-19. We observed greater overlap between dead and survivors’ parameter/biomarker distributions in the Indian cohort than in Wuhan. It was observed that KS distance between distributions of WR and IR for neutrophil and lymphocyte percentages is comparatively high while the distance between the distributions of the dead (WD, ID) across the cohorts was low. This shows that the increased overlap in the distributions in the Indian cohort is primarily due to survivors. Patients in India recovered even when their neutrophil and lymphocyte percentages reached levels similar to the levels of patients who died in Wuhan. While the observed differences could be due to better healthcare or a less severe immune response, a probable reason for the low mortality in the Delhi cohort may be the inclusion of steroids and immunosuppressant drugs in the treatment protocols by the Government of India early on in the timeline of the pandemic. Studies have shown that use of steroids like Dexamethasone lowered COVID-19 fatalities significantly when administered to patients who require supplemental oxygen [28–31]. We observed a relation between the usage of these drugs and the survival of patients with extreme lymphocytes and neutrophils counts, which are associated with mortality(Figure 8) [14, 15, 24, 32, 33].

Machine learning models for risk stratification and mortality prediction were developed based on features extracted from Indian cohort. The important features for risk stratification included blood parameters, diabetes comorbid condition and oxygen saturation level. On the other hand, mortality prediction is dependent only on blood parameters. Blood coagulation parameters (ferritin, D-Dimer and INR), immune and inflammation parameters (IL6, LDH and Neutrophil(%)) are common features for both risk and mortality prediction. Features for mortality prediction also included NLR, WBC and Trop I. Some of these features have been identified as predictors of the progression of the COVID-19 disease [12, 14, 15, 24, 33–39].

The best performing model for risk stratification on the Indian dataset was the XGboost classifier, which acheived an F1-score of 0.81±0.01 while Logistic regression yielded the best performance for mortality prediction with an F1-score of 0.71±0.02. We also examined the performance of these algorithms when trained on a dataset comprising of only vitals and clinical attributes, as these are features that can be acquired quickly and may aid in the initial decision-making process. The best performing models gave an F1 score of 0.76±0.02 for risk stratification and 0.34±0.3 for mortality prediction. The low performance of these models shows the importance of blood parameters in describing the progression of COVID-19.

We observed that the progression of COVID-19 infection is accompanied by hemocytometric changes with respect to the numbers of days to outcome (Figures 9,10). The final day of outcome was considered as it is a more stable reference point compared to the day of admission as a patient may be identified and admitted late in the progression of the disease. The patients who died showed elevated levels of D-Dimer, Ferritin and NLR, while lymphocyte (%) levels dropped. The separation of the biomarkers’ values between the two classes is observed to be consistent through the course of the disease. This shows their significance in making predictions. Interestingly, the mortality prediction model performed better when nearing the day of outcome whereas the performance of the risk stratification model decreased as we move towards the day of the outcome. The differences between the survivors and the dead increase as the time progresses as survivors recover from the conditions whereas patients who die do not, making it easier for any predictive model to classify. The performance of risk stratification decreases as we move towards the day outcome because as patients recover the differences between low risk and high-risk candidates converge, making it more difficult for the model to classify. The proposed models are based on the data collected from the Delhi region in India. This may introduce regional biases and therefore, needed to be tested across multi-center. Our study provides a preliminary assessment of the clinical course and outcome of Delhi patients. We intend to test these models in the future on larger data collected from multi-hospitals located in different geographic locations in India. As more data becomes available, the whole procedure can easily be repeated to obtain better models and more insights. Although we had a pool of about 70 clinical measurements, here our modelling principle is a trade-off between the minimal number of features and the capacity for good prediction, therefore avoiding overfitting. Nevertheless, studies done on other cohorts have also identified these features as key predictors [33].

**Figure 9.**
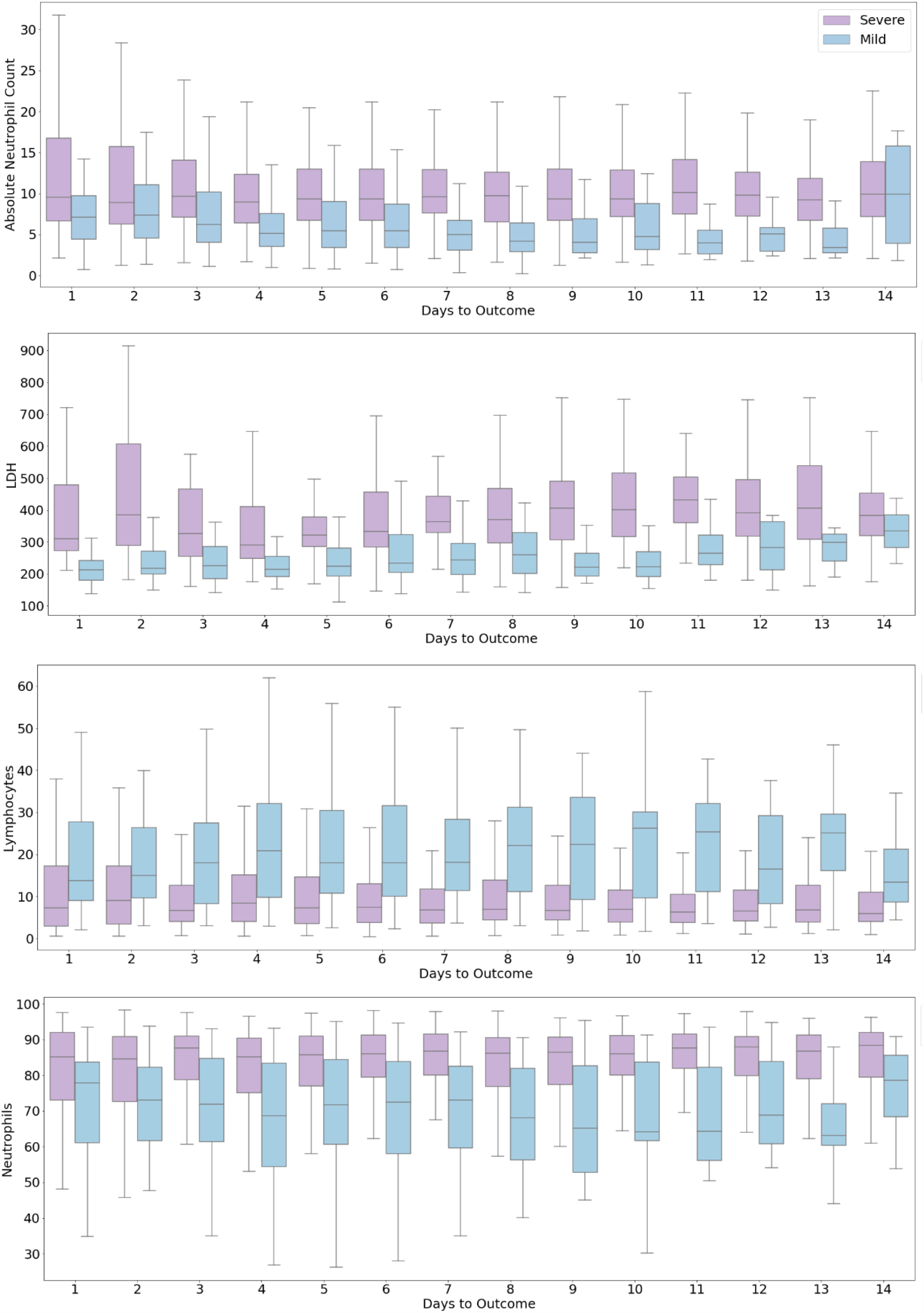
Progression of biomarkers by risk

**Figure 10.**
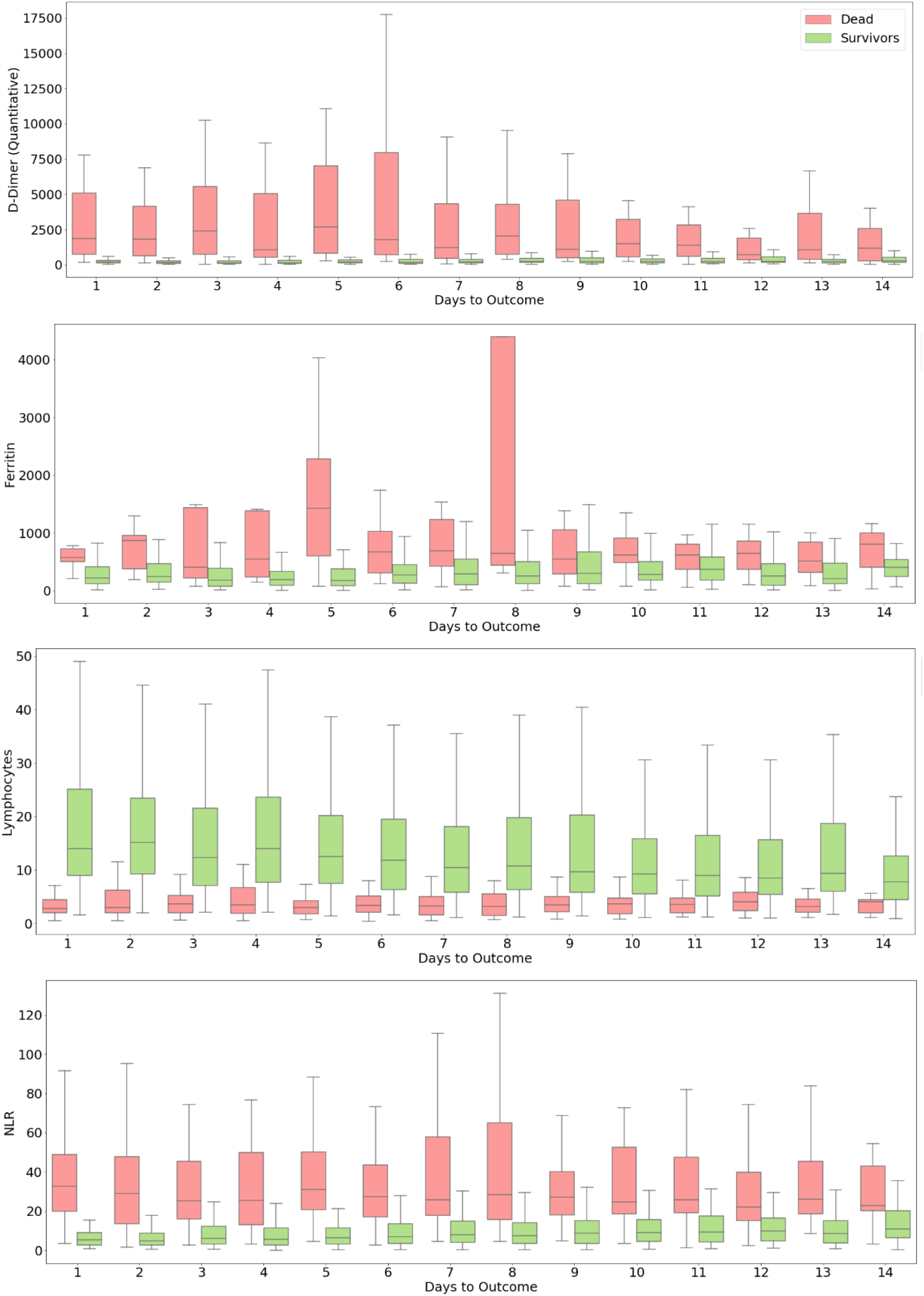
Progression of biomarkers by mortality

The major strength of our study is the inclusion of a relatively large group of confirmed COVID-19 cases from India. The findings from this study will not only help in clinical decision-making in Indian healthcare setting but will also help healthcare systems worldwide with understanding of progression of severity and the role of steroids in patient survival. This study enables to move in the direction of building accurate risk and mortality prediction models and in identifying significant trends in clinical course and in exploring the impact of individual steroids on COVID-19 patients.

## Conclusion

Accurate risk stratification and mortality prediction models based on vitals, comorbidities and blood parameters will help in rapid screening of infected patients and hence in optimal use of the healthcare infrastructure. It is likely that cohort-specific difference may emerge due to the difference in demographic conditions and healthcare setting. This necessitates the development of population specific solutions. There is also a need to study the effectiveness of certain treatment protocols affecting mortality. Our study presents the first data collection effort to develop predictive models and to study feature differences and the effect of steroids in the Indian population. Risk stratification and mortality prediction models yielded good performance and F1-scores of 0.81 and 0.71 respectively. Haematological parameters are important features for risk stratification and mortality prediction models. The analysis showed that steroids might have played a role in patient survival with extreme neutrophils or lymphocytes. This may indicate the effectiveness of the use of steroids in managing COVID19, and possibly explain the effectiveness of the treatment protocols being followed by the Indian medical systems. This study would help accelerate the decision-making process in healthcare systems for focused and efficient medical treatments.

## Data Availability

The datasets analyzed and generated during the current study would be made public once the manuscript is accepted.

## Acknowledgement

This COVID project was funded by Intel Corp as part of its Pandemic Response Technology Initiative (PRTI) [40]. The Project from IGIB side was funded by CSIR (MLP-2005) and Fondation Botnar (CLP-0031). Authors also acknowledge Dr. Mitali Mukerji for facilitating collaboration with the clinical partner. PKV and UDP also thank DST-SERB for support (CVD/2020/000343).

