## Supplementary Material for "COVID-19 Risk Stratification and Mortality Prediction in Hospitalized Indian Patients"

**Table S1.** Categorical features for Risk Stratification. Medians and P-values are given for individual features.

| Statistical Analysis for Categorical Features | | | |
|---|---|---|---|
| **Feature** | **Risk Stratification** | | |
| | **High Risk [n (%)]** | **Low Risk [n (%)]** | **P-value** |
| Sex, Female | 78 (26.0) | 86 (28.67) | 0.0508 |
| Pregnancy | 0 (0.0) | 1 (0.33) | 0.2675 |
| Steroids prescribed | 225 (75.0) | 113 (37.67) | 0.0 |
| AntiVirals prescribed | 207 (69.0) | 181 (60.33) | 0.4768 |
| Hypertension | 159 (53.0) | 89 (29.67) | 0.0045 |
| Diabetes | 147 (49.0) | 81 (27.0) | 0.0046 |
| Cancer | 7 (2.33) | 7 (2.33) | 0.6986 |
| Hyperlipidemia/Dislipidemia | 10 (3.33) | 5 (1.67) | 0.3697 |
| Thyroid related illness | 42 (14.0) | 44 (14.67) | 0.2394 |
| Heart/Circulatory System related illness | 47 (15.67) | 18 (6.0) | 0.0054 |
| Respiratory illness | 31 (10.33) | 24 (8.0) | 0.856 |
| Brain/Nervous System illness | 16 (5.33) | 8 (2.67) | 0.2565 |
| Renal illness | 28 (9.33) | 16 (5.33) | 0.2575 |
| Liver related illness | 5 (1.67) | 4 (1.33) | 0.9803 |
| presence of any Other illness | 60 (20.0) | 29 (9.67) | 0.02 |

**Table S2.** Continuous features for Risk Stratification. Medians and P-values are given for individual features.

| Statistical Analysis (Numerical Features) | | | |
|---|---|---|---|
| **Feature Name** | **Risk Stratification** | | |
| | **High Risk (IQR)** | **Low Risk (IQR)** | **P-value** |
| Glycosylated Haemoglobin(Hb A1c) (%) | 6.7 (5.94-7.7) | 6.1 (5.54-7.19) | 0.0004 |
| Average Glucose Value For the Last 3 Months (mg/dl) | 145.5 (123.7-174.2) | 128.3 (112.3-159.6) | 0.0004 |
| Average Glucose Value For the Last 3 Months IFCC (mmol/L) | 8.06 (6.86-9.65) | 7.11 (6.22-8.84) | 0.0004 |
| Glycosylated Haemoglobin(Hb A1c) IFCC (mmol/mol) | 49.72 (41.41-60.65) | 43.16 (37.04-55.07) | 0.0004 |
| Temperature (°F) | 98.2 (97.8-98.6) | 98.0 (97.3-98.6) | 0.3588 |
| BP Systolic (mmHg) | 130.0 (120.0-140.0) | 130.0 (120.0-139.0) | $\ll$0.0001 |
| BP Diastolic (mmHg) | 80.0 (70.0-84.0) | 80.0 (70.0-82.0) | $\ll$0.0001 |
| Pulse Rate | 88.0 (80.0-102.0) | 86.0 (79.0-96.0) | $\ll$0.0001 |
| $SPO_2$ (Room Air) | 94.0 (88.0-97.0) | 97.0 (96.0-98.0) | $\ll$0.0001 |
| Respiration Rate | 20.0 (20.0-24.0) | 20.0 (20.0-22.0) | $\ll$0.0001 |
| 25 Hydroxy- Vitamin D Serum (ng/mL) | 26.25 (12.94-41.76) | 22.72 (10.56-38.94) | 0.1248 |

| | | | |
|---|---|---|---|
| Basophils (%) | 0.3 (0.1-0.5) | 0.3 (0.12-0.5) | 0.8718 |
| Eosinophils (%) | 0.1 (0.0-0.5) | 0.3 (0.0-1.3) | $\ll$0.0001 |
| Haemoglobin (g/dl) | 12.2 (10.5-13.5) | 12.8 (11.0-14.0) | $\ll$0.0001 |
| Lymphocytes (%) | 6.6 (3.6-12.5) | 18.05 (9.12-29.87) | $\ll$0.0001 |
| MCH (pg) | 28.7 (26.6-30.2) | 28.9 (27.2-30.5) | 0.002 |
| MCHC (g/dl) | 33.1 (32.2-34.0) | 33.4 (32.6-34.1) | 0.0001 |
| MCV (fL) | 86.2 (81.38-90.12) | 86.4 (82.5-89.98) | 0.0319 |
| Monocytes (%) | 5.1 (3.4-7.8) | 8.0 (5.22-9.9) | $\ll$0.0001 |
| Neutrophils (%) | 87.2 (78.35-91.4) | 71.75 (59.12-84.1) | $\ll$0.0001 |
| Packed Cell Volume (%) | 36.9 (32.1-40.55) | 38.1 (33.32-41.48) | $\ll$0.0001 |
| Platelet Count ($10^9$/L) | 231.0 (165.0-311.0) | 220.5 (170.0-292.0) | 0.0187 |
| RDW (%) | 15.1 (14.1-16.5) | 14.6 (13.8-15.6) | $\ll$0.0001 |
| Total Leucocyte Count (TLC) ($10^9$/L) | 11.6 (8.7-15.85) | 7.8 (5.7-11.4) | 0.0001 |
| Absolute Lymphocyte Count ($10^9$/L) | 0.78 (0.47-1.3) | 1.36 (0.86-1.92) | $\ll$0.0001 |
| Absolute Neutrophil Count ($10^9$/L) | 9.82 (6.92-13.97) | 5.51 (3.4-9.25) | $\ll$0.0001 |
| Absolute Monocyte Count ($10^9$/L) | 0.58 (0.39-0.86) | 0.55 (0.4-0.76) | 0.0086 |
| RBC Count ($10^{12}$/L) | 4.36 (3.85-4.76) | 4.43 (3.99-4.79) | 0.0254 |
| MPV (fL) | 9.1 (8.4-10.2) | 9.2 (8.4-10.68) | 0.0129 |
| WBC | 11.59 (8.69-15.85) | 7.8 (5.7-11.4) | 0.0001 |
| NLR | 13.19 (6.33-25.24) | 3.94 (1.99-8.82) | $\ll$0.0001 |
| LMR | 1.39 (0.84-2.15) | 2.42 (1.56-3.56) | $\ll$0.0001 |
| NMR | 16.59 (9.99-26.95) | 8.54 (6.3-15.39) | $\ll$0.0001 |
| PLR | 300.0 (177.7-525.0) | 169.9 (109.6-268.1) | $\ll$0.0001 |
| CRP (mg/L) | 23.66 (6.37-72.56) | 8.99 (2.99-32.71) | $\ll$0.0001 |
| Absolute Basophil Count ($10^9$/L) | 0.04 (0.02-0.07) | 0.03 (0.02-0.05) | $\ll$0.0001 |
| Absolute Eosinophil Count ($10^9$/L) | 0.05 (0.02-0.12) | 0.06 (0.02-0.12) | 0.3466 |
| Ferritin (ng/mL) | 387.2 (211.0-706.2) | 147.4 (66.1-302.0) | $\ll$0.0001 |
| Trop I (ng/mL) | 0.01 (0.01-0.03) | 0.01 (0.0-0.01) | 0.1159 |
| Procalcitonin Level (ng/mL) | 0.11 (0.07-0.26) | 0.05 (0.03-0.1) | 0.0044 |
| CK-MB (Mass) (ng/mL) | 2.0 (1.0-4.7) | 1.1 (0.7-1.9) | 0.0194 |
| IL-6 (pg/ml) | 35.94 (9.73-122.25) | 7.84 (3.25-22.43) | $\ll$0.0001 |
| INR | 1.13 (1.04-1.27) | 1.05 (0.99-1.16) | 0.0554 |
| Prothrombin Time (sec) | 12.8 (11.8-14.4) | 12.0 (11.2-13.1) | 0.0596 |
| D-Dimer (Quantitative) (ng/mL) | 384.0 (188.0-1122.9) | 143.5 (81.3-239.9) | $\ll$0.0001 |
| Magnesium (mg/dl) | 2.1 (1.9-2.3) | 2.0 (1.9-2.2) | 0.0208 |
| LDH (IU/L) | 384.5 (301.0-508.2) | 231.0 (193.0-292.0) | $\ll$0.0001 |
| Creatine Kinase (U/L) | 106.0 (52.5-209.25) | 86.5 (60.75-154.25) | 0.0542 |
| E (%) | 24.76 (21.38-27.9) | 24.13 (20.83-27.07) | 0.2 |
| RdRp (mg/dl) | 27.15 (22.2-29.36) | 25.35 (22.82-28.16) | 0.56 |
| Age (yrs) | 61.0 (53.0-68.0) | 53.0 (41.0-64.0) | $\ll$0.0001 |
| Height (cms) | 162.0 (157.0-170.0) | 165.0 (158.0-171.2) | 0.8566 |
| Weight (kg) | 73.15 (63.08-82.0) | 72.5 (63.0-80.0) | 0.4477 |
| BMI | 26.32 (23.89-30.11) | 26.31 (24.05-28.13) | 0.108 |

**Table S3.** Categorical features for Mortality Prediction. Medians and P-values are given for individual features.

| Statistical Analysis for Categorical Features | | | |
|---|---|---|---|
| **Feature** | **Risk Stratification** | | |
| | **Died [n (%)]** | **Survived [n (%)]** | **P-value** |
| Sex, Female | 16 (26.23) | 148 (30.64) | 0.55 |
| Pregnancy | 0 (0.0) | 1 (0.21) | 0.72 |
| Steroids prescribed | 35 (57.38) | 303 (62.73) | 0.62 |
| AntiVirals prescribed | 31 (50.82) | 357 (73.91) | 0.04 |
| Hypertension | 40 (65.57) | 208 (43.06) | 0.01 |
| Diabetes | 34 (55.74) | 194 (40.17) | 0.08 |
| Cancer | 3 (4.92) | 11 (2.28) | 0.23 |
| Hyperlipidemia/Dislipidemia | 0 (0.00) | 15 (3.11) | 0.17 |
| Thyroid related illness | 10 (16.39) | 76 (15.73) | 0.9 |
| Heart/Circulatory System related illness | 14 (22.95) | 51 (10.56) | 0.01 |
| Respiratory illness | 7 (11.48) | 48 (9.94) | 0.72 |
| Brain/Nervous System illness | 3 (4.92) | 21 (4.35) | 0.84 |
| Renal illness | 12.0 (19.67) | 32.0 (6.63) | 0.0 |
| Liver related illness | 3.0 (4.92) | 6.0 (1.24) | 0.04 |
| presence of any Other illness | 15.0 (24.59) | 74.0 (15.32) | 0.09 |

**Table S4.** Continuous features for Mortality Prediction. Medians and P-values are given for individual features.

| Statistical Analysis (Numerical Features) | | | |
|---|---|---|---|
| **Feature Name** | **Mortality Prediction** | | |
| | **Died (IQR)** | **Survived (IQR)** | **P-value** |
| Glycosylated Haemoglobin(Hb A1c) (%) | 6.17 (5.76-7.12) | 6.38 (5.7-7.52) | 0.3997 |
| Average Glucose Value For the Last 3 Months (mg/dl) | 130.38 (118.61-157.5) | 136.26 (116.96-169.05) | 0.3997 |
| Average Glucose Value For the Last 3 Months IFCC (mmol/L) | 7.22 (6.57-8.72) | 7.54 (6.48-9.36) | 0.3998 |
| Glycosylated Haemoglobin(Hb A1c) IFCC (mmol/mol) | 43.92 (39.44-54.25) | 46.16 (38.81-58.65) | 0.3997 |
| Temperature (°F) | 98.2 (98.0-98.6) | 98.1 (97.5-98.6) | 0.0038 |
| BP Systolic (mmHg) | 130.0 (124.0-140.0) | 130.0 (120.0-140.0) | 0.0862 |
| BP Diastolic (mmHg) | 80.0 (70.0-80.0) | 80.0 (70.0-84.0) | $\ll$0.0001 |
| Pulse Rate | 88.0 (80.0-107.0) | 88.0 (80.0-100.0) | $\ll$0.0001 |
| $SPO_2$ (Room Air) | 92.0 (80.0-96.0) | 96.0 (93.0-98.0) | $\ll$0.0001 |
| Respiration Rate | 22.0 (20.0-26.0) | 20.0 (20.0-22.0) | $\ll$0.0001 |

| | | | |
|---|---|---|---|
| 25 Hydroxy- Vitamin D Serum (ng/mL) | 26.06 (14.52-39.53) | 24.1 (11.07-40.64) | 0.494 |
| Basophils (%) | 0.2 (0.1-0.4) | 0.3 (0.2-0.6) | ≪0.0001 |
| Eosinophils (%) | 0.1 (0.0-0.3) | 0.1 (0.0-0.8) | ≪0.0001 |
| Haemoglobin (g/dl) | 11.5 (9.7-13.3) | 12.5 (10.9-13.7) | ≪0.0001 |
| Lymphocytes (%) | 3.8 (2.0-5.6) | 10.1 (5.55-18.8) | ≪0.0001 |
| MCH (pg) | 28.7 (26.7-30.2) | 28.7 (26.8-30.3) | 0.4111 |
| MCHC (g/dl) | 32.8 (31.8-33.7) | 33.3 (32.4-34.1) | ≪0.0001 |
| MCV (fL) | 87.0 (82.18-90.7) | 86.1 (81.4-90.0) | ≪0.0001 |
| Monocytes (%) | 3.9 (2.6-5.9) | 6.3 (4.0-8.9) | ≪0.0001 |
| Neutrophils (%) | 91.0 (87.45-93.8) | 82.4 (70.8-89.2) | ≪0.0001 |
| Packed Cell Volume (%) | 35.4 (30.05-39.4) | 37.5 (33.1-41.0) | ≪0.0001 |
| Platelet Count ($10^9$/L) | 185.0 (155.0-265.5) | 241.0 (174.0-319.0) | ≪0.0001 |
| RDW (%) | 15.3 (14.4-16.7) | 14.9 (14.0-16.2) | 0.0001 |
| Total Leucocyte Count (TLC) ($10^9$/L) | 15.5 (10.9-20.8) | 10.2 (7.3-13.3) | ≪0.0001 |
| Absolute Lymphocyte Count ($10^9$/L) | 0.56 (0.31-0.9) | 1.0 (0.6-1.64) | 0.0479 |
| Absolute Neutrophil Count ($10^9$/L) | 13.99 (9.62-18.9) | 8.19 (5.24-11.29) | ≪0.0001 |
| Absolute Monocyte Count ($10^9$/L) | 0.56 (0.37-0.91) | 0.58 (0.4-0.83) | ≪0.0001 |
| RBC Count ($10^{12}$/L) | 4.16 (3.5-4.66) | 4.43 (3.97-4.79) | ≪0.0001 |
| MPV (fL) | 9.6 (8.65-10.5) | 9.0 (8.3-10.1) | ≪0.0001 |
| WBC | 15.5 (10.9-20.8) | 10.2 (7.3-13.3) | ≪0.0001 |
| NLR | 23.97 (15.65-45.44) | 8.15 (3.78-15.95) | ≪0.0001 |
| LMR | 0.9 (0.55-1.52) | 1.73 (1.14-2.67) | ≪0.0001 |
| NMR | 23.57 (14.97-35.83) | 12.72 (7.93-21.85) | ≪0.0001 |
| PLR | 385.71 (207.61-691.49) | 242.73 (145.91-419.21) | ≪0.0001 |
| CRP (mg/L) | 59.01 (18.33-122.56) | 14.7 (4.3-50.36) | ≪0.0001 |
| Absolute Basophil Count ($10^9$/L) | 0.04 (0.02-0.08) | 0.03 (0.02-0.06) | ≪0.0001 |
| Absolute Eosinophil Count ($10^9$/L) | 0.04 (0.02-0.12) | 0.05 (0.02-0.12) | 0.0224 |
| Ferritin (ng/mL) | 603.9 (327.75-1052.4) | 260.0 (119.85-505.65) | ≪0.0001 |
| Trop I (ng/mL) | 0.02 (0.01-0.14) | 0.01 (0.0-0.01) | ≪0.0001 |
| Procalcitonin Level (ng/mL) | 0.18 (0.09-0.67) | 0.08 (0.05-0.16) | 0.0055 |
| CK-MB (Mass) (ng/mL) | 3.9 (1.6-8.3) | 1.3 (0.8-2.7) | ≪0.0001 |
| IL-6 (pg/ml) | 157.1 (39.27-626.9) | 16.13 (4.94-51.61) | ≪0.0001 |
| INR | 1.26 (1.14-1.52) | 1.1 (1.02-1.19) | ≪0.0001 |
| Prothrombin Time (sec) | 14.3 (12.95-17.3) | 12.4 (11.5-13.55) | ≪0.0001 |
| D-Dimer (Quantitative) (ng/mL) | 1386.5 (488.75-3987.55) | 219.0 (126.5-454.5) | ≪0.0001 |
| Magnesium (mg/dl) | 2.1 (1.9-2.48) | 2.1 (1.9-2.2) | 0.0496 |
| LDH (IU/L) | 527.0 (404.0-743.0) | 310.0 (233.0-400.25) | ≪0.0001 |
| Creatine Kinase (U/L) | 123.0 (58.0-282.5) | 89.0 (57.0-170.0) | 0.1142 |

| | | | |
|---|---|---|---|
| E (%) | 22.94 (15.72-27.69) | 24.76 (21.48-27.67) | ≪0.0001 |
| RdRp (mg/dl) | 26.98 (18.24-28.45) | 26.5 (22.82-29.36) | ≪0.0001 |
| Age (years) | 65.5 (60.75-70.25) | 57.0 (46.0-65.5) | ≪0.0001 |
| Height (cms) | 160.2 (159.5-167.0) | 165.0 (157.0-170.0) | 0.918 |
| Weight (kg) | 65.1 (61.45-74.15) | 73.0 (63.0-82.0) | 0.2572 |
| BMI | 24.08 (23.25-25.75) | 26.63 (24.04-28.8) | 0.1646 |

**Table S5.** Performance of the developed machine learning algorithms in risk stratification reported as mean ± standard deviation.

| **Risk Stratification** | | | |
|---|---|---|---|
| **Algorithm** | **AUC** | **F1 score** | **Average Precision** |
| XGBoost | 0.833 ± 0.01 | 0.810 ± 0.01 | 0.891 ± 0.01 |
| Random forest | 0.826 ± 0.01 | 0.803 ± 0.01 | 0.878 ± 0.01 |
| SVM | 0.817 ± 0.01 | 0.752 ± 0.01 | 0.889 ± 0.01 |
| Logistic regression | 0.812 ± 0.01 | 0.759 ± 0.01 | 0.885 ± 0.01 |

**Table S6.** Performance of the developed machine learning algorithms in mortality prediction reported as mean ± standard deviation.

| **Mortality Prediction** | | | |
|---|---|---|---|
| **Algorithm** | **AUC** | **F1 score** | **Average Precision** |
| XGBoost | 0.891 ± 0.02 | 0.634 ± 0.02 | 0.732 ± 0.03 |
| Random forest | 0.858 ± 0.01 | 0.555 ± 0.03 | 0.615 ± 0.03 |
| SVM | 0.895 ± 0.01 | 0.659 ± 0.02 | 0.710 ± 0.02 |
| Logistic regression | 0.927 ± 0.01 | 0.710 ± 0.02 | 0.801 ± 0.02 |

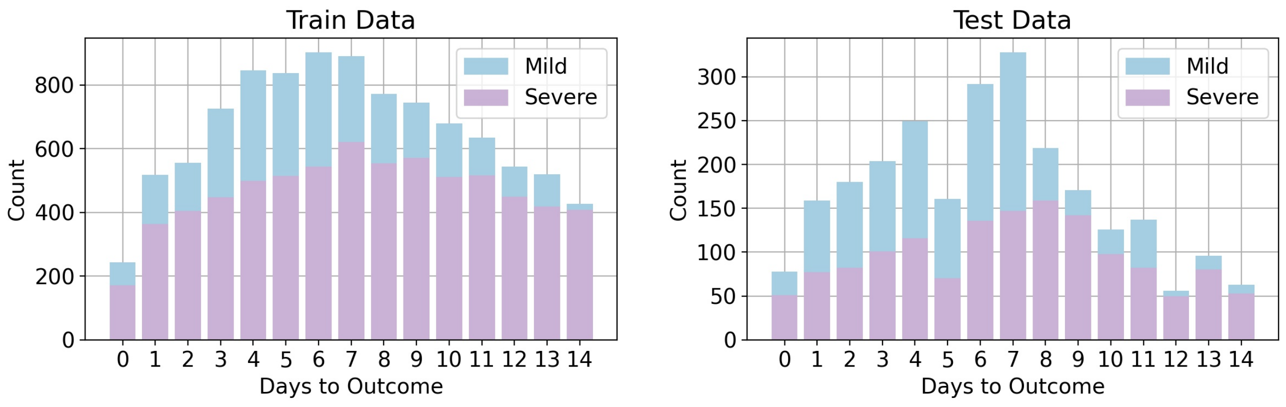


**Figure S1.** Sample Distribution for Risk Stratification with time to outcome

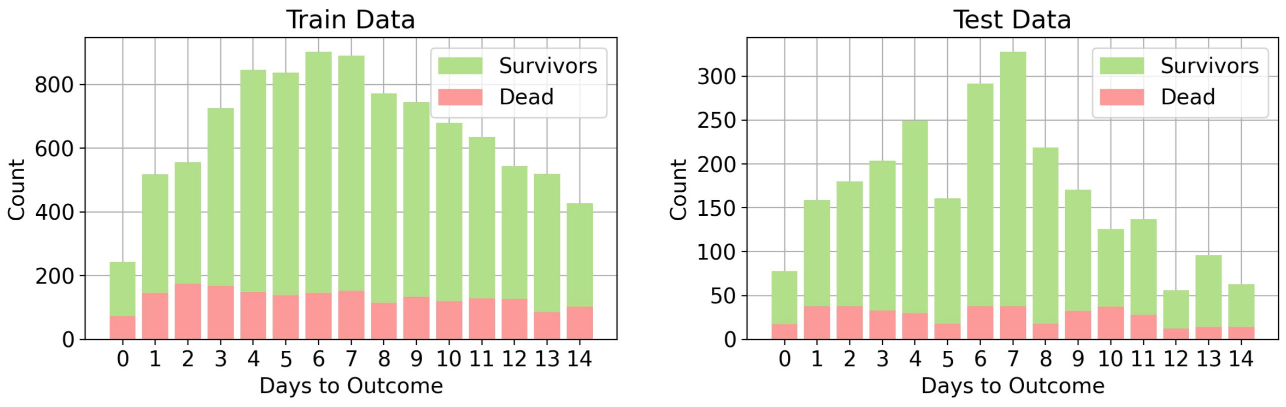


**Figure S2.** Sample Distribution for Mortality Prediction with time to outcome

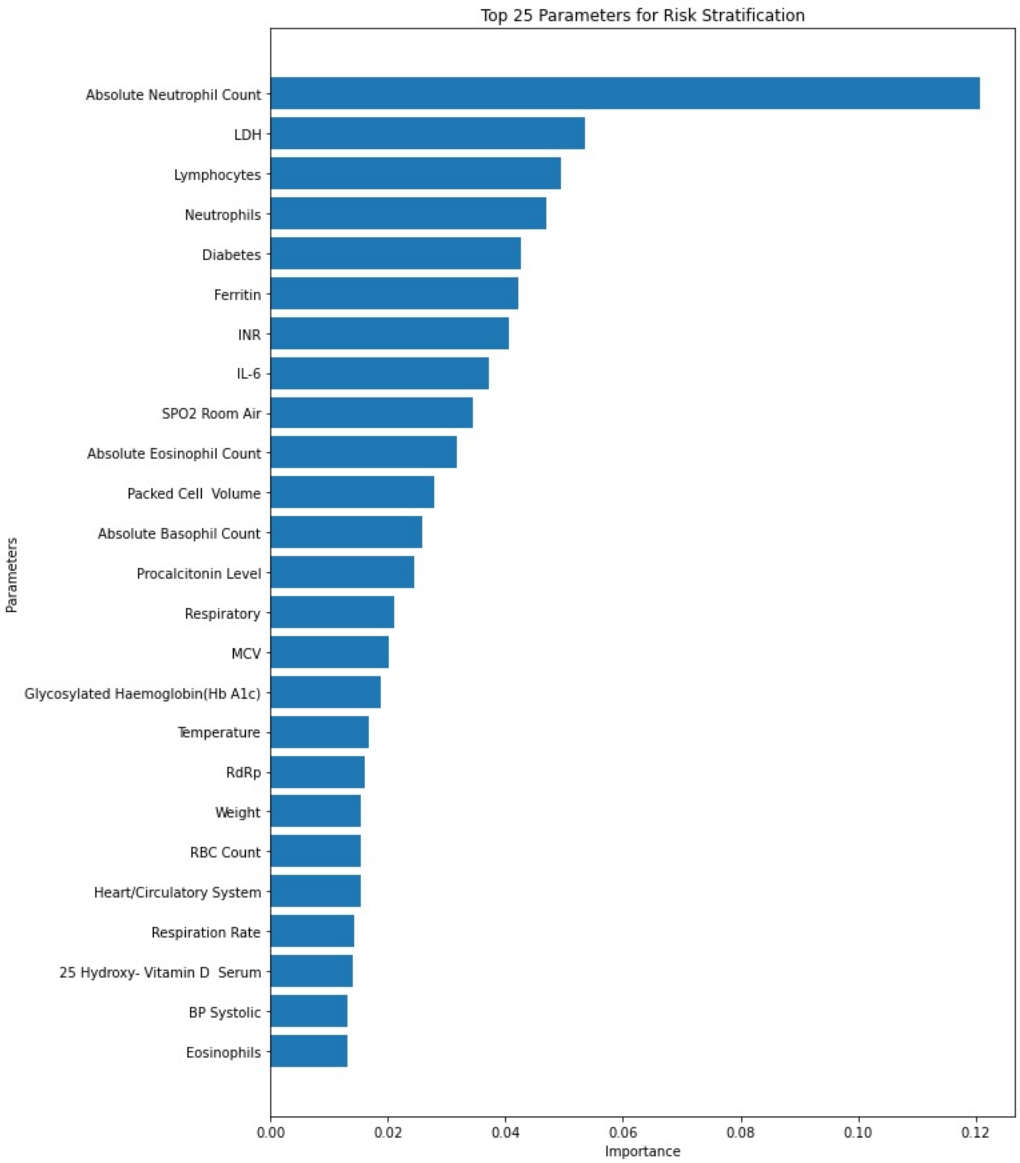


**Figure S3.** Top 25 Important Parameters for Binary Risk Stratification

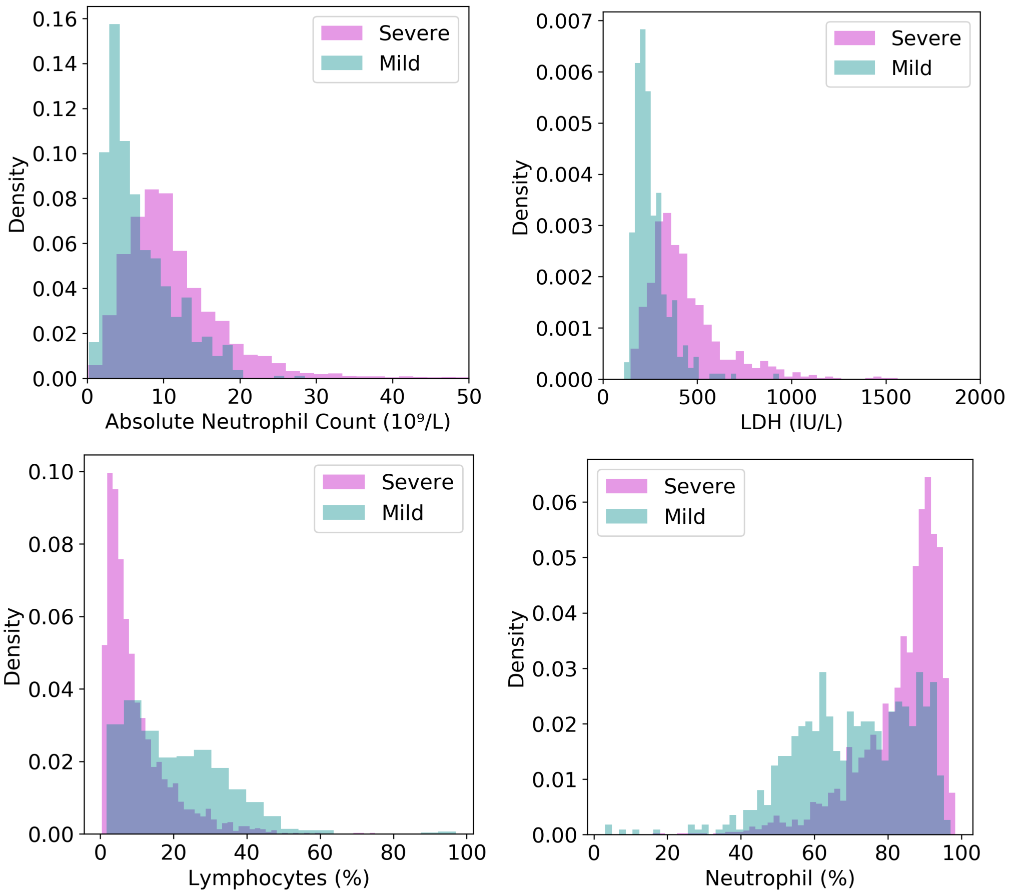


**Figure S4.** Distribution plots for four most important features used for risk stratification.

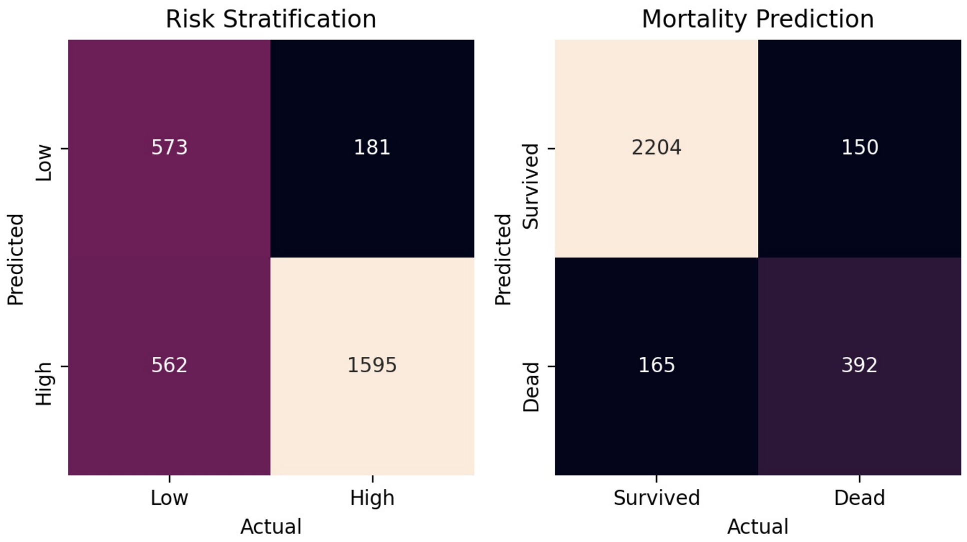


**Figure S5.** Confusion Matrices for Mortality Prediction with logistic regression and Risk Stratification with XGBoost Classifier trained on entire train set

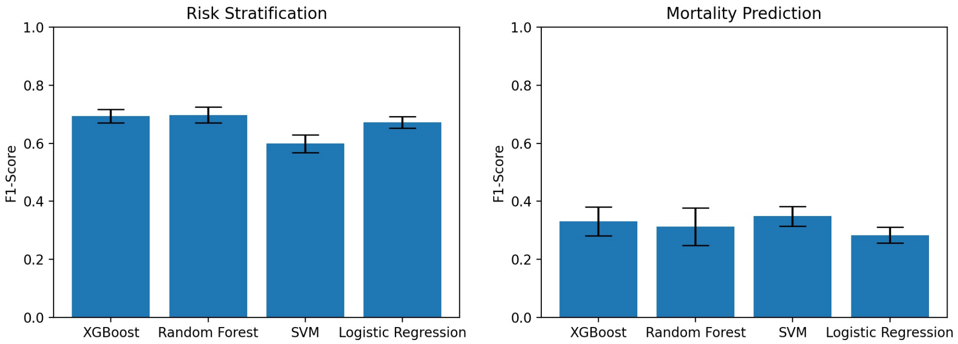


**Figure S6.** Comparison of F1 scores for various machine learning models that use only patient vitals

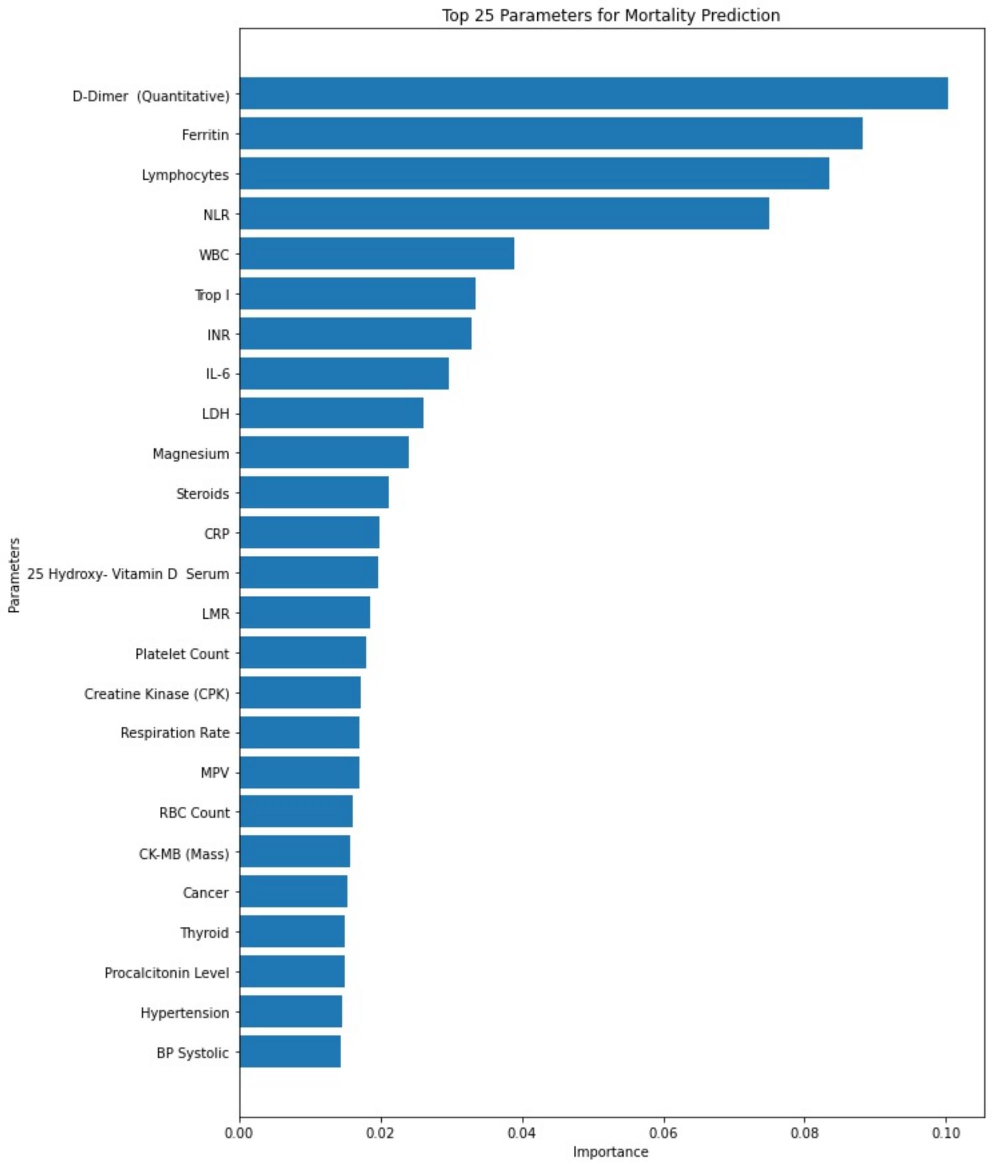


**Figure S7.** Top 25 Important Parameters for Mortality prediction

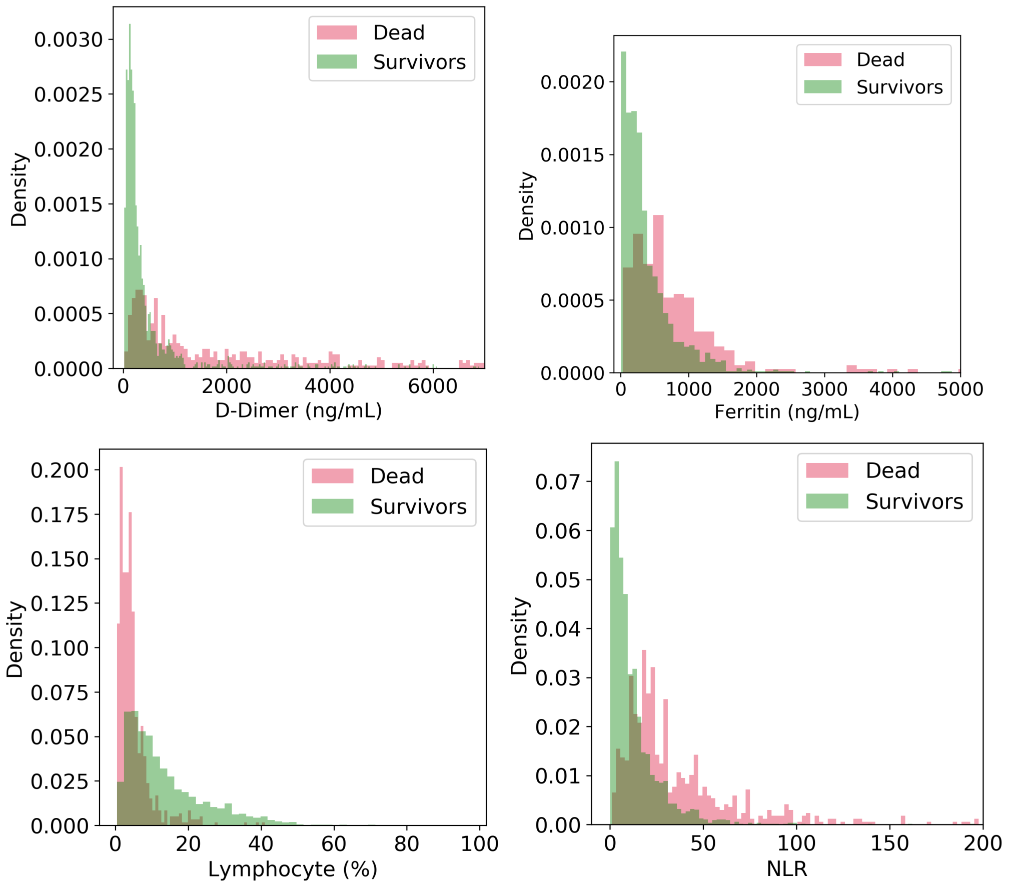


**Figure S8.** Distribution plots for four most important features used for predicting mortality.
